# The association of personality traits with childhood obesity: a systematic review and meta-analysis

**DOI:** 10.1101/2022.06.22.22276748

**Authors:** Dong-Xia Jiang, Tian-Yu Huang, Jing Chen, Wu-Cai Xiao, Zheng Liu

## Abstract

A growing body of evidence has shown the association of personality with obesity, but findings of such studies in children remain mixed. This review aimed to systematically summarize the existing literature regarding the association between personality and childhood obesity and explore the underlying mechanisms. A meta-analysis was performed based on the data from 12 included studies and personality traits were classified into five domains (Extraversion, Agreeableness, Conscientiousness, Neuroticism, and Openness) based on the five-factor model. Conscientiousness was significantly associated with BMI across 2 studies and the pooled correlation coefficient was −0.09 (95% CI: −0.17 to 0.00; I^2^ = 0%); another 3 studies indicated a significant difference in Conscientiousness between children with obesity and those with normal weight, and the pooled SMD was −0.08 (95% CI: −0.13 to −0.03; I^2^ = 66%). No consistent patterns were found in the associations between other personality traits and childhood obesity. We hypothesized three possible pathways (moderation, mediation, and confounding) underlying the personality-obesity associations. Findings of our review indicate that, within the context of childhood obesity interventions, identification of subgroups of children with specific personality traits is necessary, especially for measures of Conscientiousness.

## Introduction

Childhood obesity has become a global health concern.^1^ The prevalence of obesity among US children aged 2 to 19 years was 17.8% in 2013–2016.^2^ This figure in several developing countries has risen to 7.0%–14.4% and is predicted to increase further in coming years.^3^ Childhood obesity affects the normal growth and development during childhood and its harm to health will continue to adulthood, increasing the risk of hypertension, diabetes, coronary heart disease, and other cardiovascular diseases. Therefore, it is vital to combat childhood obesity.

Childhood obesity is a multifactorial disease influenced by the interplay of genetic, behavioral, psychological, and environmental factors. Of note, less has been explored about whether stable psychological factors possibly relate to the risk of childhood obesity. Personality, as a sum of an individual’s diverse psychological characteristics, is the enduring set of traits and styles that he or she exhibits, which characteristics represent natural tendencies of this person, and ways in which this person differs from the “standard normal person” in his or her society.^4^ Personality is a complex construct, and the five-factor model (FFM) has been widely used to guide the analysis of the construction of personality due to its good validity across different cultures and age groups of children.^5–7^ The FFM comprises five major domains: Extraversion (the tendency to be sociable, self-confident, and outgoing), Agreeableness (the tendency to be altruistic and emotionally supportive), Conscientiousness (the tendency to be self-controlled, perseverance, and disciplined to social norms), Neuroticism (the tendency to be nervousness, unstableness, and stressful), and Openness (the tendency to be imaginative, curious, and creative).^7–9^

Several lines of evidence have indicated that personality traits are closely related to one’s health behaviors and thus may be associated with the risk of obesity. However, studies exploring the associations between personality traits and obesity have led to heterogeneous findings.^9–18^ Characteristics of the study population, measures of exposures and outcomes, study design, and risk of bias may be responsible for the heterogeneity. For example, a longitudinal study among 5,265 adults aged 25-65 years in Australia observed that only Extraversion was positively associated with weight gain at 2-year follow up, while a cross-sectional study in a sample of 2,928 children aged 4–12 years living in the United States found that children with overweight scored higher on Openness and Agreeableness whereas those with obesity scored lower on these two traits than their peers with normal weight.^19,20^

To the best of our knowledge, previous reviews about the association of personality traits with obesity have been mostly focused on the adult population with one exception, which was published in 2004 and only focused on one dimension of the FFM.^9,10,18,21^ Notably, personality traits such as Conscientiousness tend to increase along the life course, while obesity-prone behaviors possibly decrease with age.^21^ Thus findings from previous reviews focusing on the adults cannot directly translate to the children. To fill in the research gap, this review was to systematically evaluate the associations of personality traits with obesity and explore the underlying mechanisms in the children population.

## Methods

We registered the protocol of the systematic review at the International Prospective Register of Systematic Reviews (PROSPERO; registration number CRD42022306529).^22^ This report follows the Preferred Reporting Items for Systematic Reviews and Meta-analyses Statement (PRISMA) 2020.^23^

### Search strategy

We searched databases of PubMed, Embase, PsycINFO, and Cochrane Central Register of Controlled Trials until May 1, 2022. Briefly, we searched literature focusing on the topics of personality, children, and obesity. Details of the search strategy are shown in the **Supporting Information**.

### Eligibility criteria

Literature that met all of the following criteria were included: (i) study subjects: children aged 3-18 years old (children under 3 years old were excluded as their personality are likely to be unstable and unmatured); (ii) study design: observational study (cross-sectional, case-control, or longitudinal study); (iii) measures of personality traits: valid and reliable questionnaires or scales; and (iv) measures of obesity: including but not limited to body mass index (BMI), BMI-z score, waist circumference (WC), general overweight/obesity (assessed by cutoffs of BMI or z score), and central obesity (assessed by cutoffs of waist-related indicators).

Literature that met any of the following criteria were excluded: (i) articles unavailable in full text; (ii) intervention study; (iii) articles not written in English or Chinese; or (iv) conference, editorial articles.

### Screening and data extraction

Literature screening was conducted at two stages. First we screened the title and abstract of each article and then assessed its full text to identify whether it met the eligibility criteria. We extracted information from the included study, including authors, publication year, country, study design, sample size, gender distribution and age, assessment of personality traits, measurement of obesity, covariates, and results. Personality traits were classified into five domains according to the FFM, including Extraversion, Agreeableness, Conscientiousness, Neuroticism, and Openness.^24^ We also summarized other personality traits such as, Self-control (or, similarly, problem in impulse control or higher reward responsiveness), Psychoticism, Persistence, and Conformity.

### Summary of measures and synthesis methods

We conducted random-effect meta-analyses to quantitatively synthesize the data using the same effect measure. The between-study heterogeneity was assessed by the I^2^ statistic. The correlation coefficients (*r*), assessing associations between personality traits and BMI, were synthesized after the Fisher’s z transformation. The mean difference, assessing whether personality traits differed between groups of children with obesity and those with normal weight, were synthesized after standardizing the effect size to a uniform scale, i.e., calculation of standardized mean difference (SMD; SMD = difference in mean outcome between groups / standard deviation of outcome among children). The odds ratios (OR) were pooled to determine the association of high/low level of personality traits with the risk of obesity.

All analyses were stratified by domains of personality traits. Studies not included in the meta-analysis were summarized qualitatively according to the Synthesis Without Meta-analysis guideline.^25^ We used R (version 4.1.2) to conduct data synthesis.

### Assessment of the study quality

We used the Newcastle-Ottawa Scale (NOS) to assess the quality of included studies.^26^ The NOS comprises 8 items within 3 categories: selection, comparability, and exposure (for the case-control or cross-sectional study)/outcome (for the cohort study). A maximum of 1 point was given for each item within the selection and exposure/outcome categories, and a maximum of 2 points was given for each item within the comparability category. A total score summing all of the points given to the 8 items ranged from 0 to 9 points. The scores of 5 or less, 6-7, and 8 or more were rated as low, medium, and high quality, respectively.

## Results

### Literature screening

The flow of literature screening is shown in **Figure 1**. There were 21 records left after eliminating the duplicates and screening the titles and abstracts. Another 7 records were identified through other sources. Finally, we assessed the full text of these 28 records and a total of 12 studies were included.

**Figure 1:**
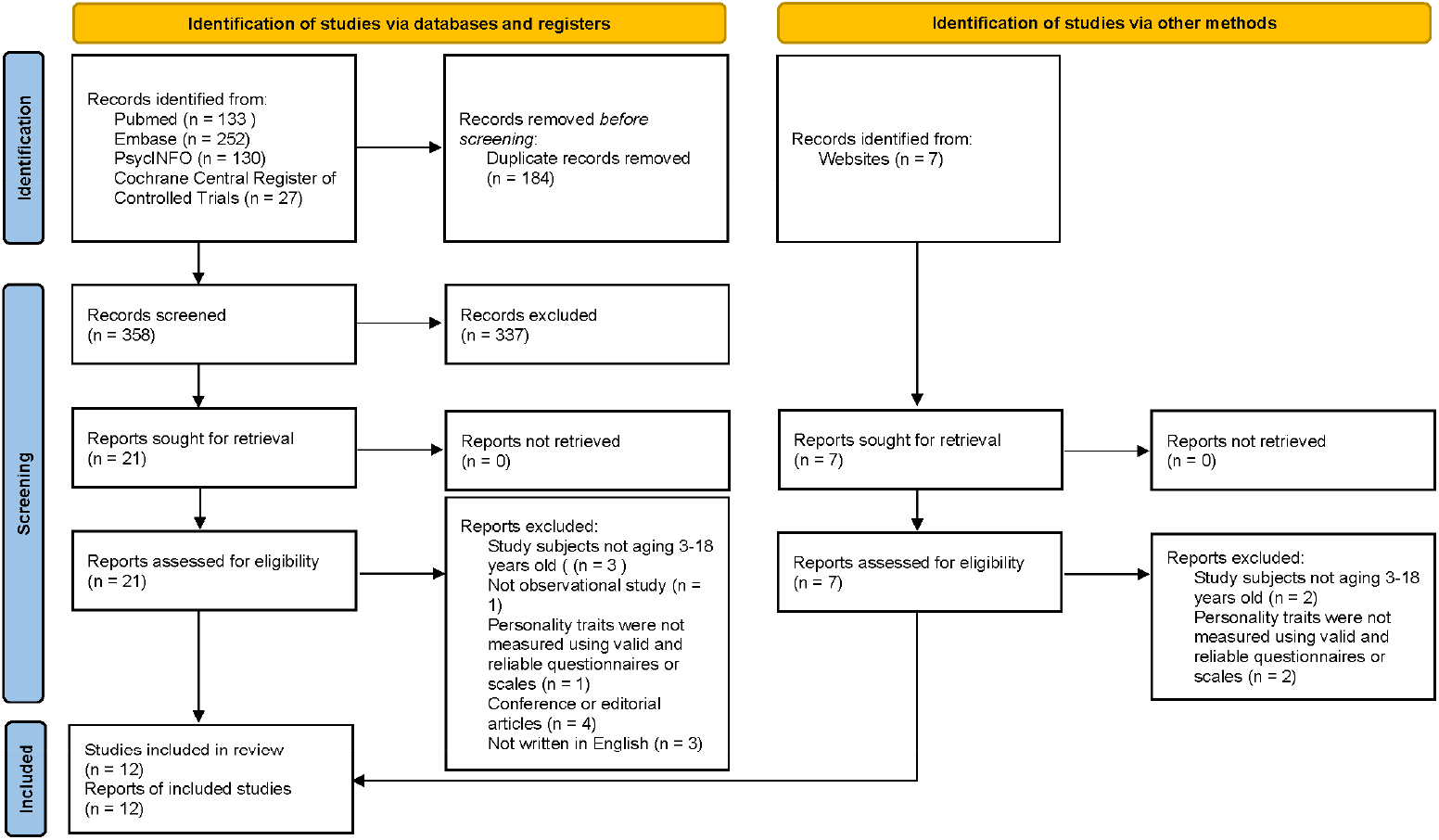
Study flow of the review

### Characteristics of included studies

Characteristics of the included 12 studies are presented in **Table 1**.^17,20,27–36^ Among the included 12 studies, 7 of them assessed Extraversion, 6 assessed Neuroticism, 5 assessed Agreeableness, Conscientiousness, and Openness, and 7 assessed other personality traits. Concerning study design, 2 of them were case-control, 5 were cross-sectional, 3 were cohort, and the remaining 2 included both cross-sectional and longitudinal samples. Sample size varied from 105 to 3857 and the percentage of girls varied from 44.2% to 61.3%. Most studies included children aged 6 to 12 years old. Four studies were conducted in America, 2 studies in Belgium, and 1 in Norway, China, Australia, Italy, Ireland, and the Netherlands, respectively.

**Table 1:**
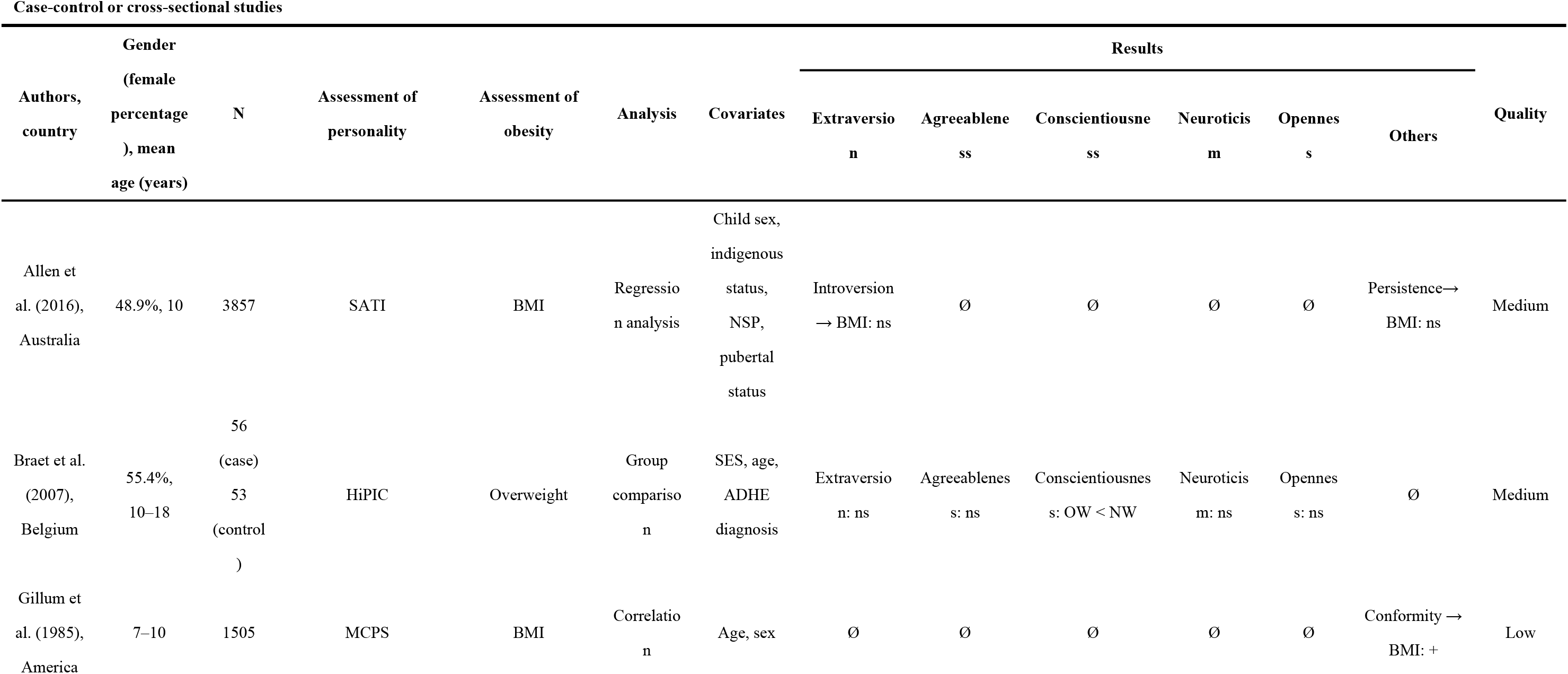
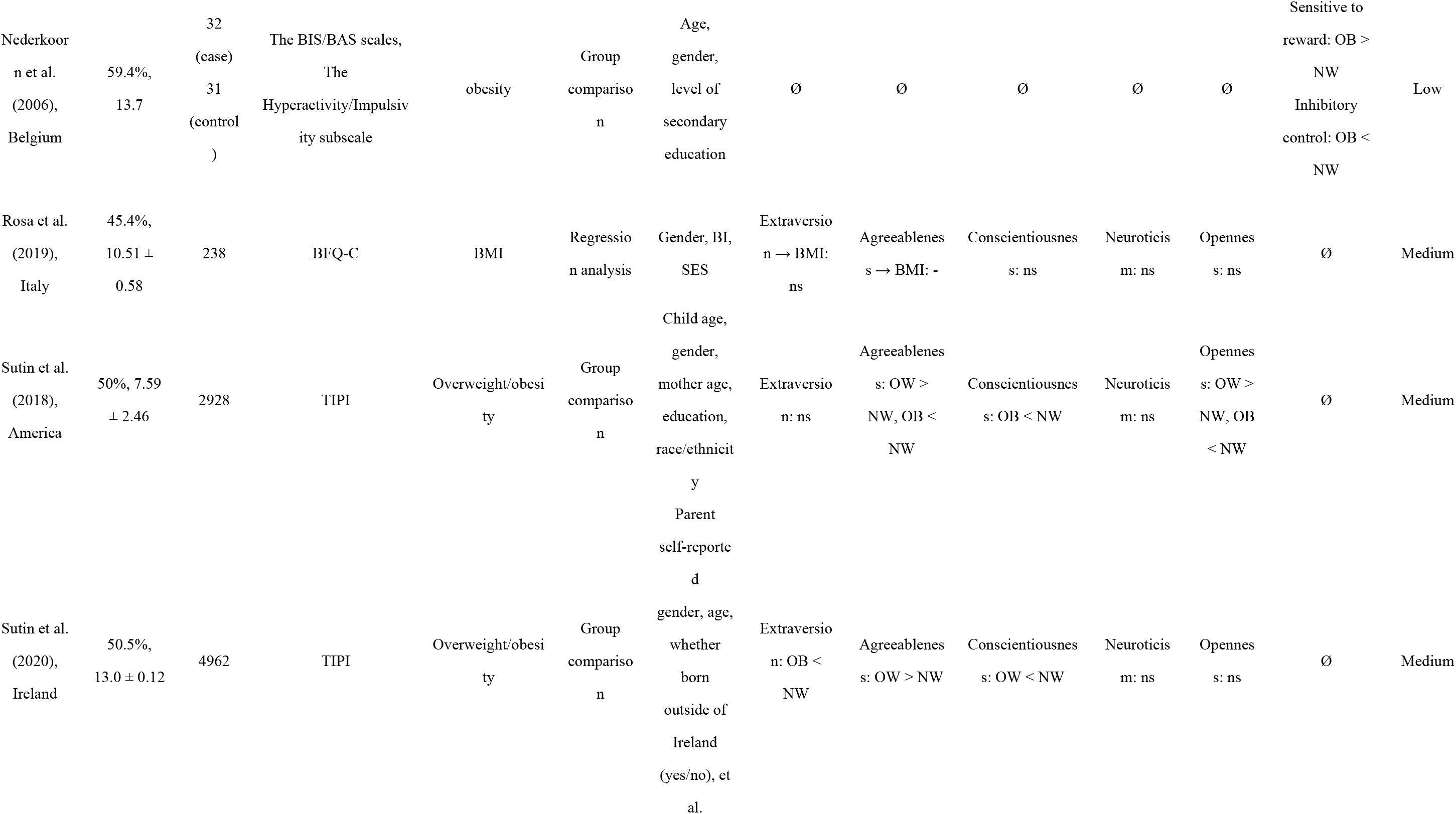
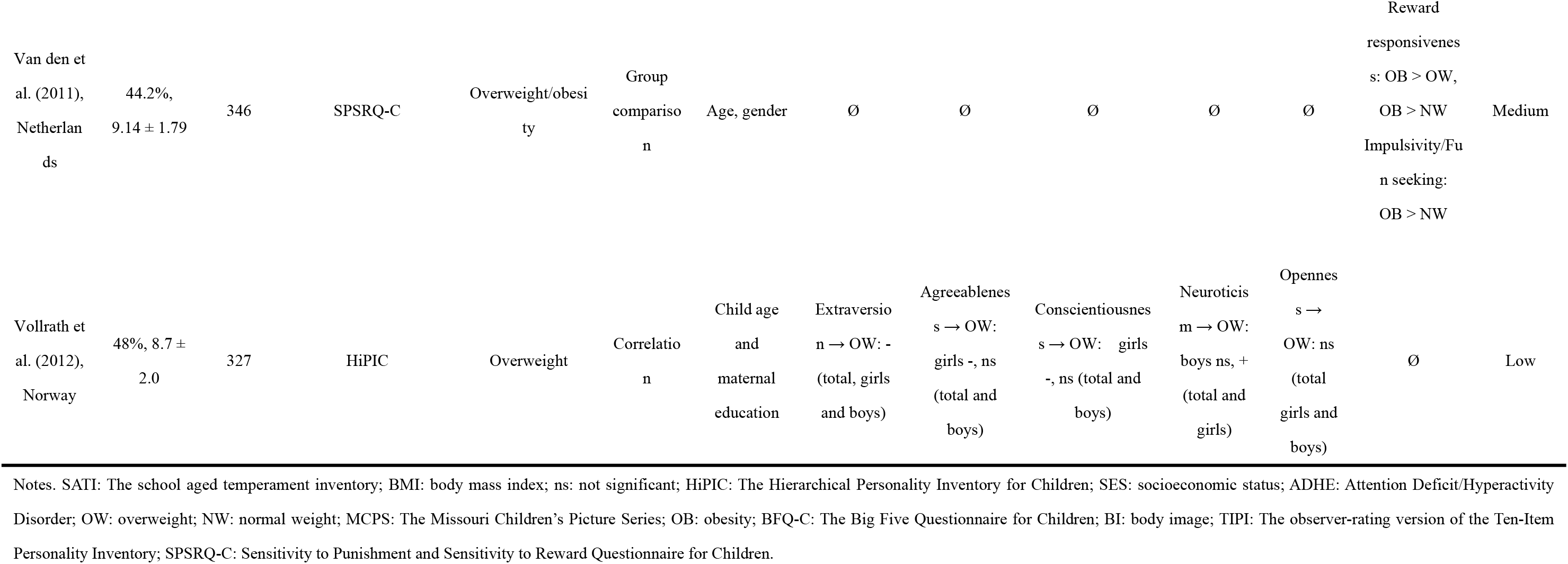
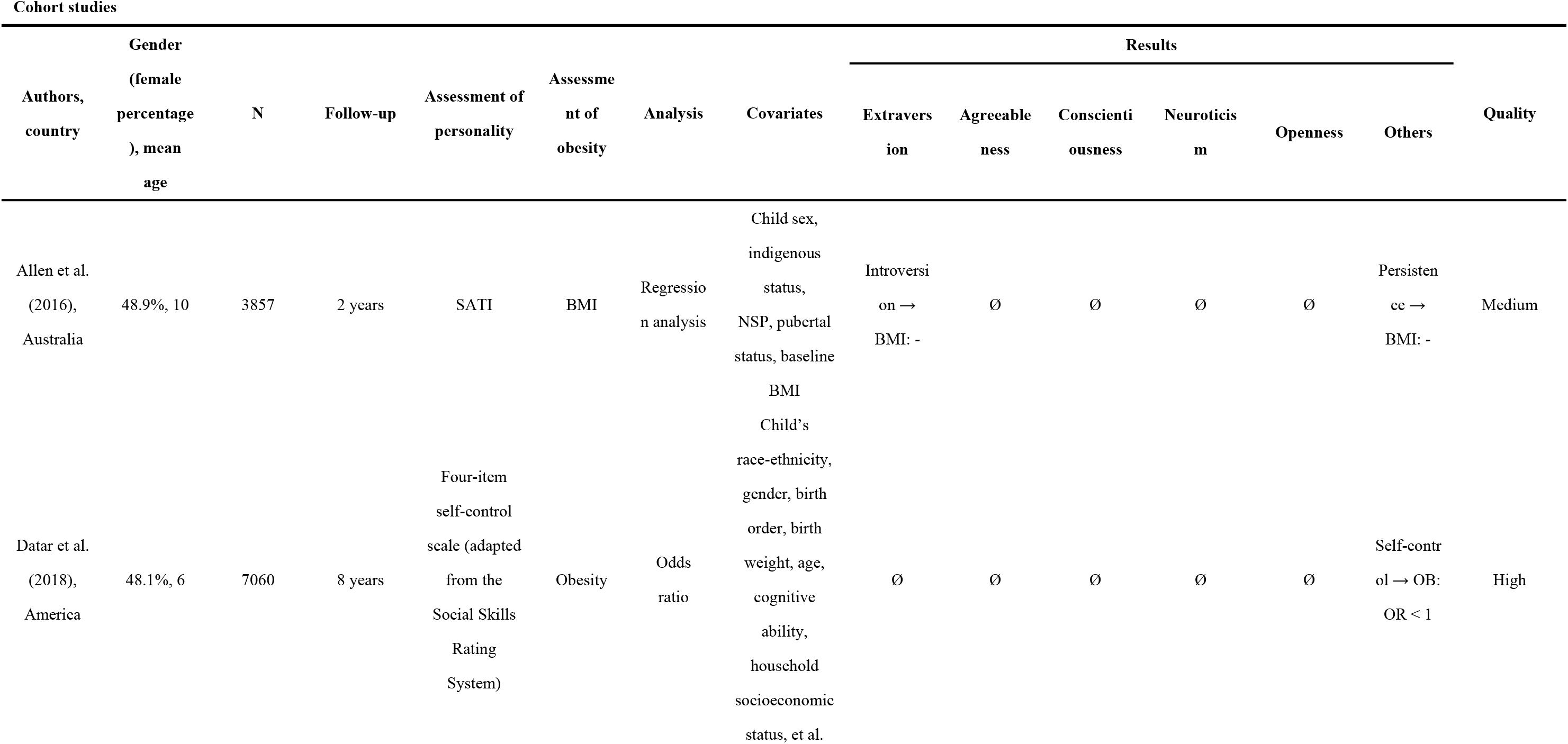
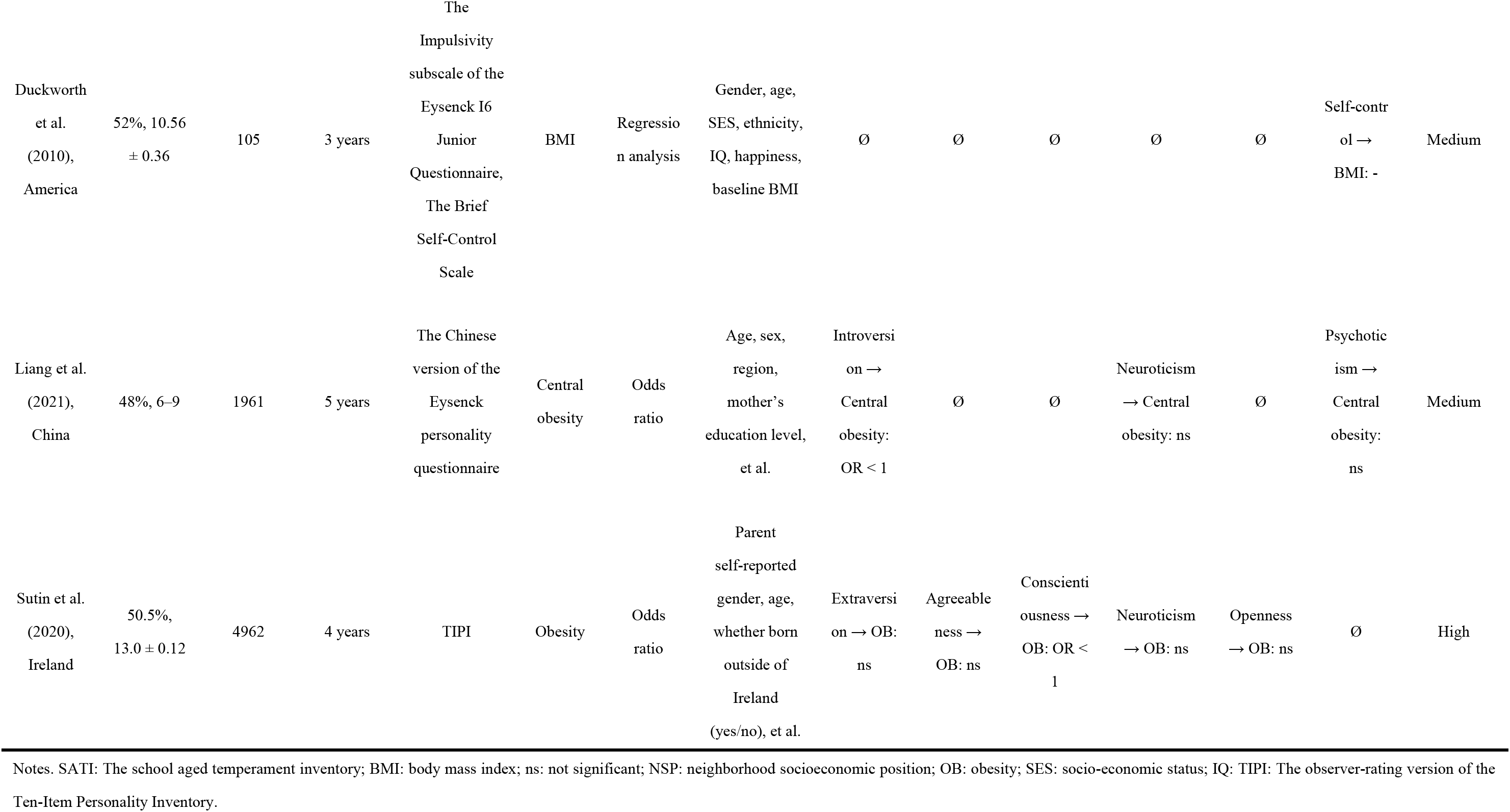
Characteristics of the included studies

### Quality of included studies

Results of quality assessment are shown in **Table 2**. Most studies were rated 3 points within the selection category, 2 points within the comparability category, and 2 points within the exposure/outcome category. There were 1, 7, and 3 studies rated as high, medium, and low quality, respectively. For a study including both cross-sectional and longitudinal analyses, the longitudinal part was rated as medium quality while the cross-sectional part as high quality.

**Table 2:**
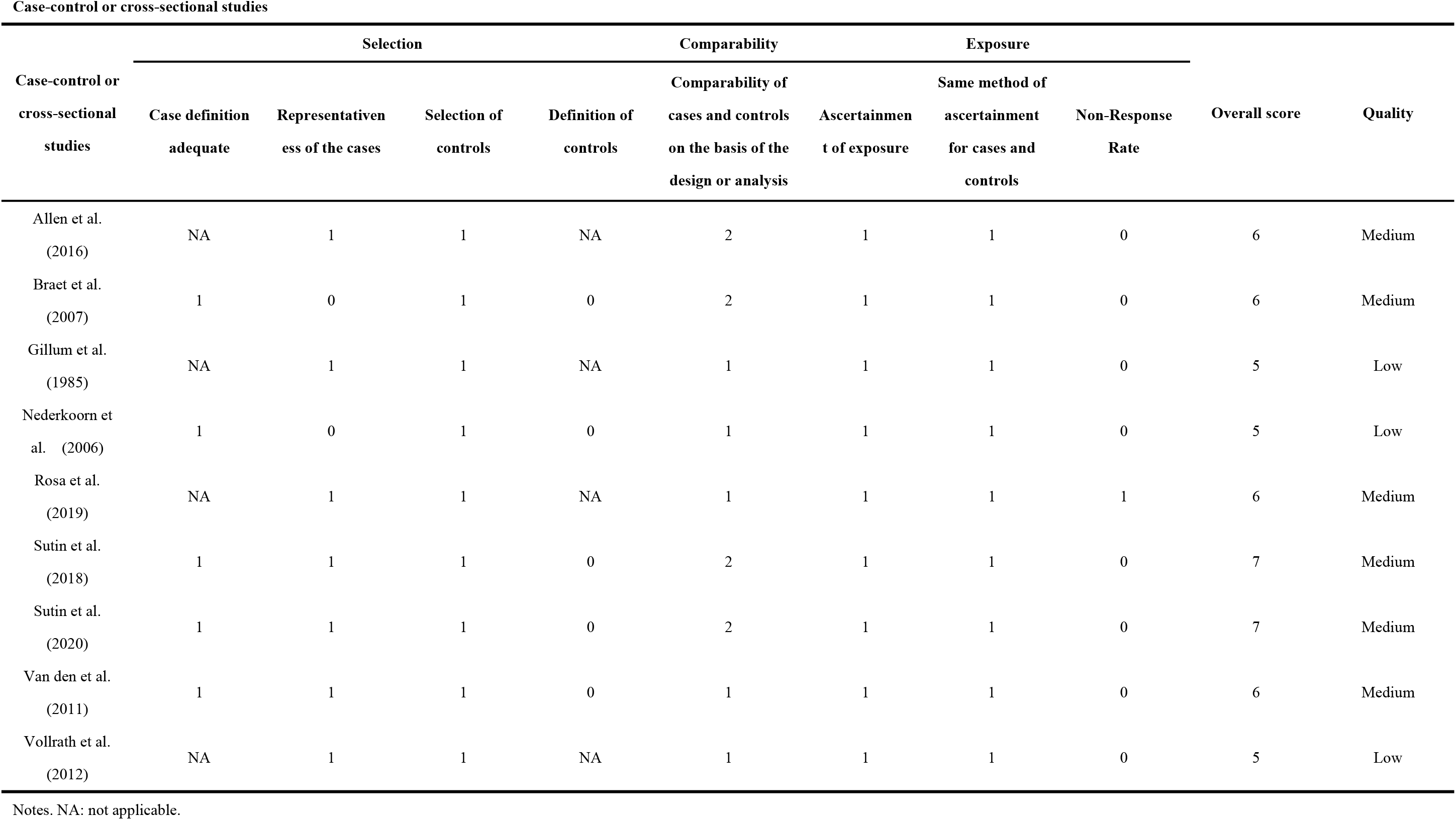
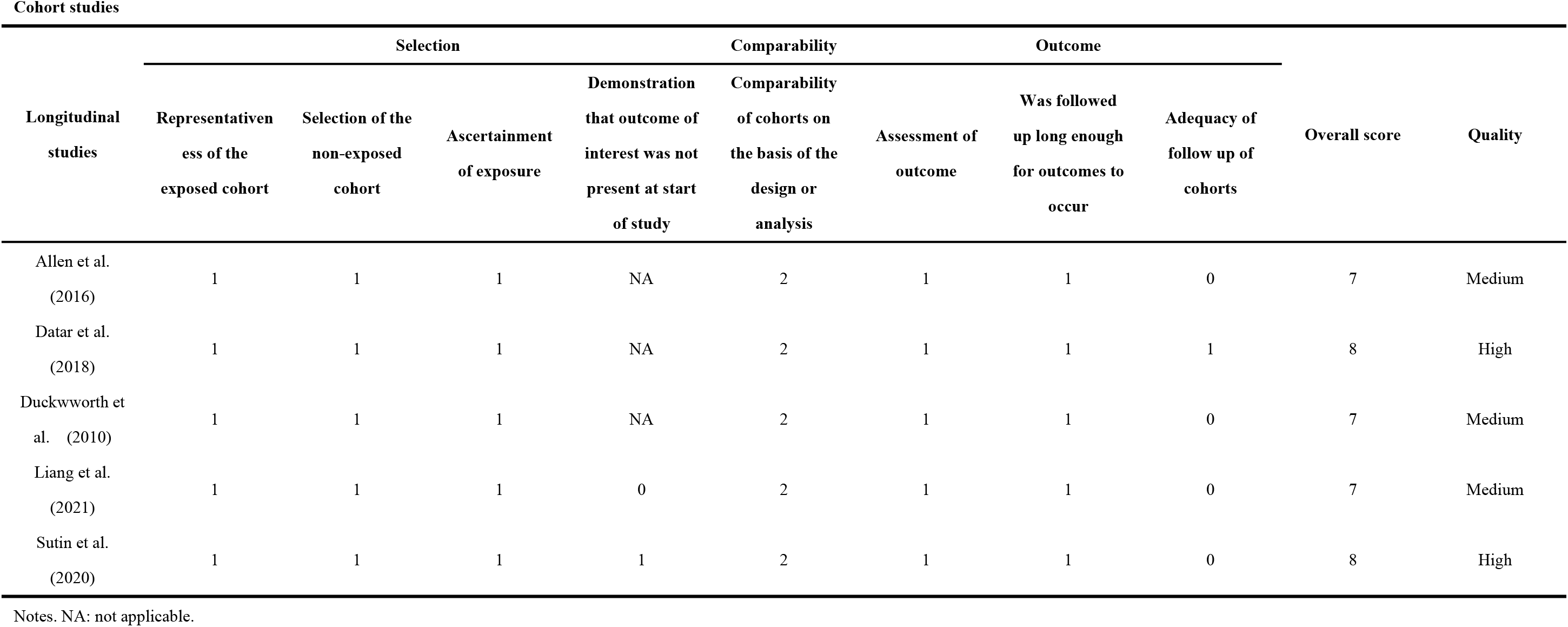
Quality of included studies

### Association between personality traits and childhood obesity

#### Association between Extraversion and childhood obesity

Among the 7 studies included for this analysis, 3 measured correlation coefficients,^27,34,36^ 3 for SMD,^20,28,35^ and 2 for OR.^32,35^ The pooled correlation coefficient was −0.02 (95% CI: −0.10 to 0.06; I^2^ = 61%) (**Figure 2**), indicating that Extraversion was not significantly associated with BMI. The pooled SMD was −0.02 (95% CI: −0.07 to 0.03; I^2^ = 0%) (**Figure 3**), indicating a non-significant difference of Extraversion between children with obesity and those with normal weight. The pooled OR was 0.99 (95% CI: 0.98 to 1.00; I^2^ = 0%) (**Figure 4**), showing that children with high level of Extraversion had a significantly but slightly lower risk of obesity.

**Figure 2:**
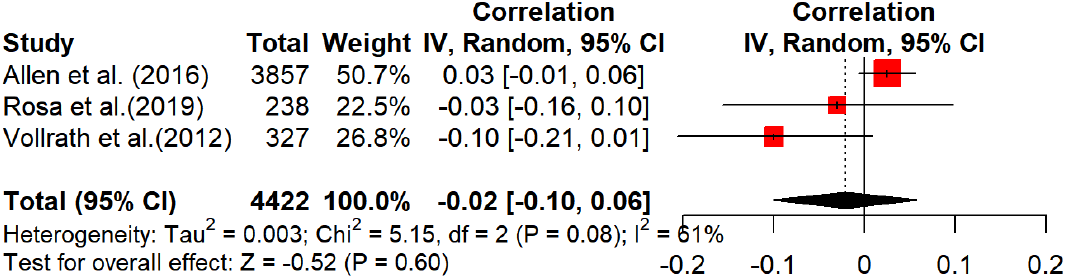
Forest plot showing the correlation coefficients between Extraversion and BMI

**Figure 3:**
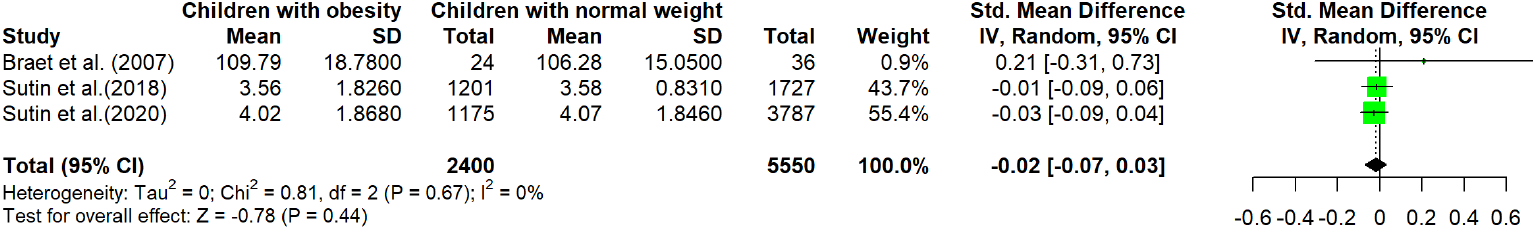
Forest plot showing the standardized mean difference of Extraversion between children with obesity and those with normal weight

**Figure 4:**
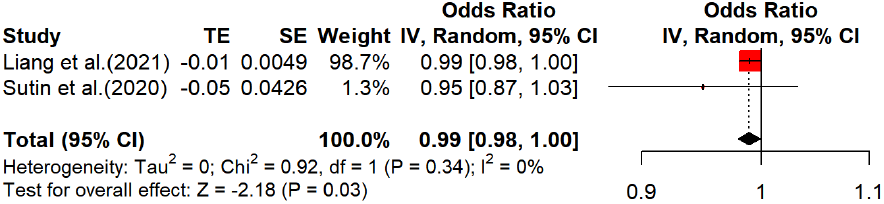
Forest plot showing the odds ratio of baseline level of Extraversion as a predictor of the follow-up risk of obesity

A longitudinal study not included in the meta-analysis due to different effect measures found that children with a lower level of Introversion (opposite to Extraversion) at baseline had a greater increase in BMI at 2-year follow up, after controlling baseline BMI, sex, indigenous status, pubertal status, and neighborhood socioeconomic position (beta coefficient = −0.12, 95% CI: −0.21 to −0.03, *P* < 0.05).^27^

#### Association between Agreeableness and childhood obesity

Among the 5 studies included for this analysis, 2 measured correlation coefficients,^34,36^ and 3 measured SMD.^20,28,35^ The pooled correlation coefficient was −0.07 (95% CI: −0.17 to −0.01; I^2^ = 0%) (**Figure 5**), suggesting that Agreeableness was significantly and negatively associated with BMI. The pooled SMD was 0.04 (95% CI: −0.06 to 0.13; I^2^ = 57%) (**Figure 6**), indicating a non-significant difference in Agreeableness between children with obesity and those with normal weight.

**Figure 5:**
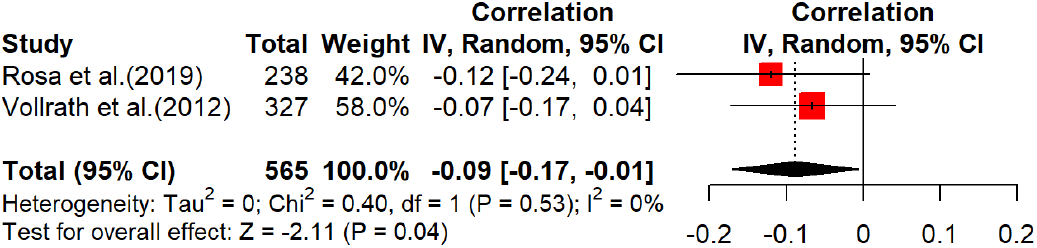
Forest plot showing the correlation coefficients between Agreeableness and BMI

**Figure 6:**
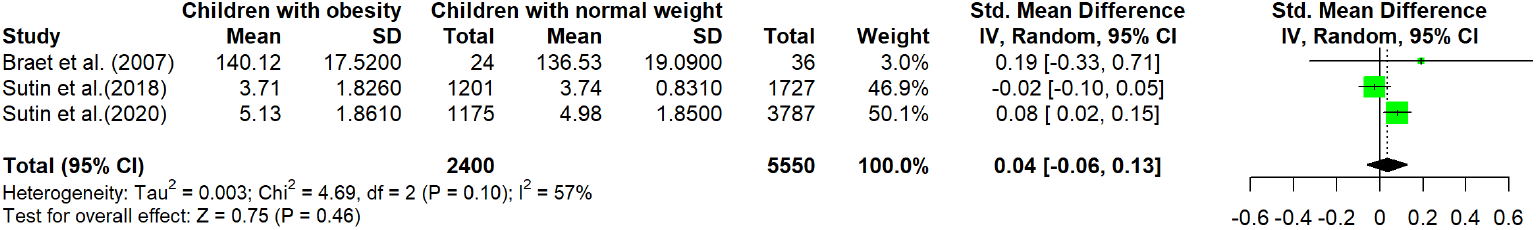
Forest plot showing the standardized mean difference of Agreeableness between children with obesity and those with normal weight

The longitudinal analysis of a study not included in the meta-analysis due to different effect measures found that there was no significant association between Agreeableness and risk of obesity over the 4-year follow-up (OR = 0.95, 95% CI: 0.88 to 1.04).^35^

#### Association between Conscientiousness and childhood obesity

Among the 5 studies included for this analysis, 2 measured correlation coefficients,^34,36^ and 3 used SMD as the effect measure.^20,28,35^ The pooled correlation coefficient was −0.09 (95% CI: −0.17 to 0.00; I^2^ = 0%) (**Figure 7**), suggesting that Conscientiousness was significantly associated with BMI. The pooled SMD was −0.08 (95% CI: −0.13 to −0.03; I^2^ = 66%) (**Figure 8**), indicating a significant difference of Conscientiousness between children with obesity and those with normal weight.

**Figure 7:**
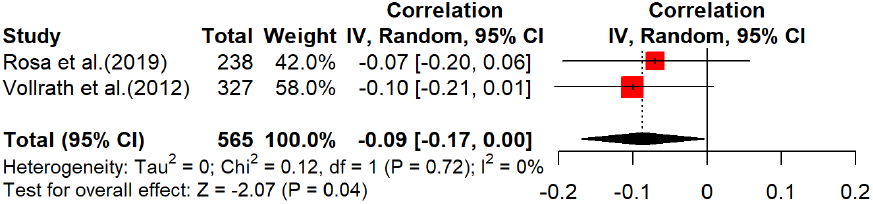
Forest plot showing the correlation coefficients between Conscientiousness and BMI

**Figure 8:**
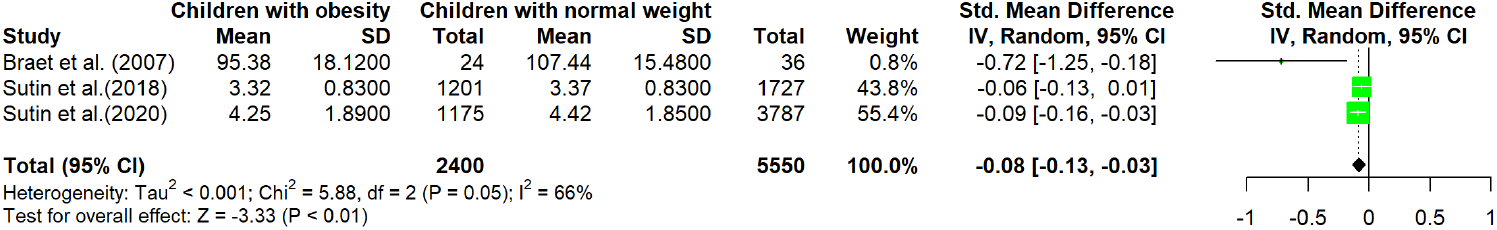
Forest plot showing the standardized mean difference of Conscientiousness between children with obesity and those with normal weight

The longitudinal analysis of a study not included in the meta-analysis due to different effect measures also found higher Conscientiousness was associated with lower risk of developing obesity over 4-year follow-up (OR = 0.88, 95% CI: 0.82 to 0.96).^35^

#### Association between Neuroticism and childhood obesity

Among the 5 studies included for this analysis, 2 measured correlation coefficients,^34,36^ and 3 for SMD.^20,28,35^ The pooled correlation coefficient was −0.01 (95%CI: −0.09 to 0.07; I^2^ = 0%) (**Figure 9**), suggesting that Neuroticism was not significantly associated with BMI. The pooled SMD was −0.02, 95%CI: −0.07 to 0.04; I^2^ = 5%) (**Figure 10**), indicating a non-significant difference in Neuroticism between children with obesity and those with normal weight.

**Figure 9:**
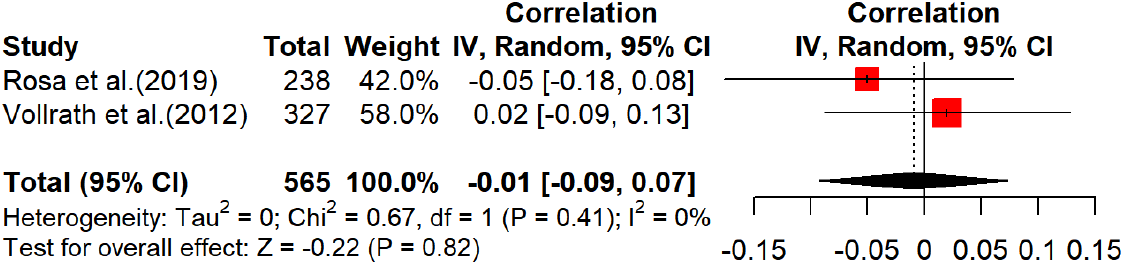
Forest plot showing the correlation coefficients between Neuroticism and BMI

**Figure 10:**
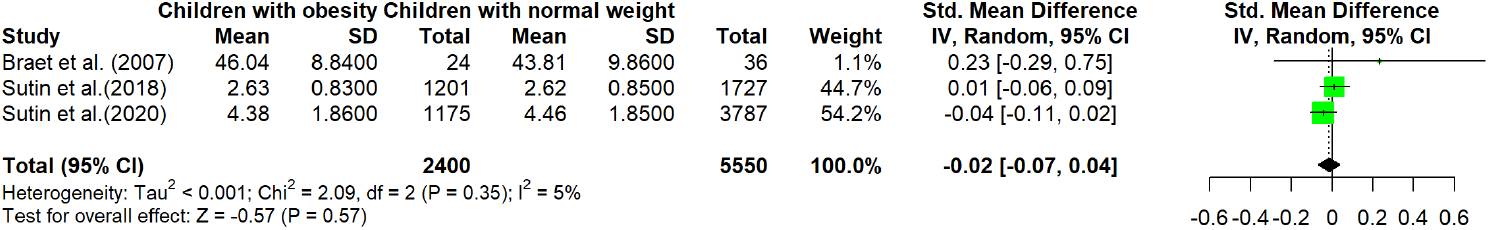
Forest plot showing the standardized mean difference of Neuroticism between children with obesity and those with normal weight

The longitudinal study by Liang et al. which was not included in the meta-analysis due to different effect measures found no significant association between Neuroticism and central obesity at 4-year follow-up in 1961 children (OR = 1.003, 95%CI: 0.994 to 1.011).^32^

#### Association between Openness and childhood obesity

Among the 5 studies included for this analysis, 2 measured correlation coefficients,^34,36^ and 3 used SMD as the effect measure.^20,28,35^ The pooled correlation coefficient was −0.09 (95%CI: −0.17 to 0.00; I^2^ = 0%) (**Figure 11**), suggesting that Openness was significantly and negatively associated with BMI. The pooled SMD was 0.05 (95%CI: −0.03 to 0.13; I^2^ = 55%) (**Figure 12**), indicating a non-significant difference in Openness between children with obesity and those with normal weight.

**Figure 11:**
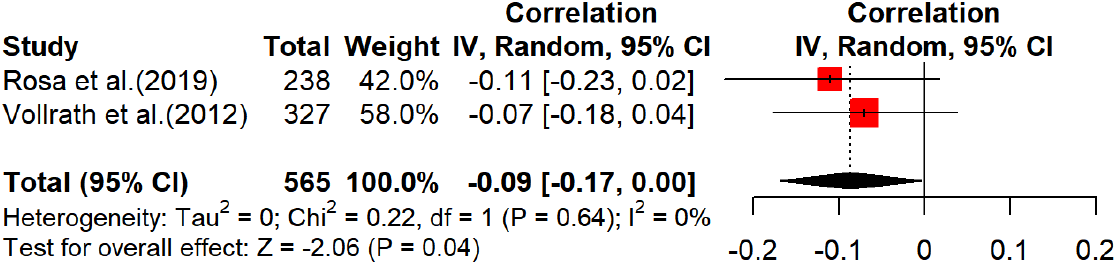
Forest plot showing the correlation coefficients between Openness and BMI

**Figure 12:**
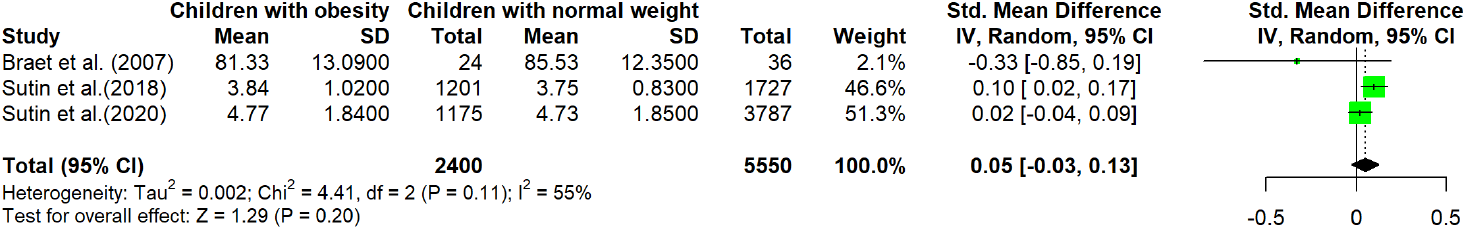
Forest plot showing the standardized mean difference of Openness between children with obesity and those with normal weight

The longitudinal analysis of a study not included in the meta-analysis due to different effect measures found that there was no significant association between Openness and risk of obesity over the 4-year follow-up (OR = 0.99, 95% CI: 0.91 to 1.08).^35^

#### Association between other personality traits with childhood obesity

Seven studies assessed the associations between personality traits other than five dimensions comprised in the FFM and childhood obesity. Four studies found that the reduced self-control (or, similarly, problem in impulse control or higher reward responsiveness) was related to higher BMI in children.^17,29,30,33^ A longitudinal study found no significant association of Psychoticism with central obesity in 1961 children.^32^ Allen et al. found that lower level of Persistence at baseline was associated with a higher BMI at 2-year follow-up.^27^ Gillum et al. assessed several dimensions of personality traits by the Missouri Children’s Picture Series in 1506 children and only the conformity (complying with social rules) score was significantly and positively correlated with BMI (r= 0.08, *P< 0.01*).^31^

## Discussion

### Summary of study findings

In this study, we systematically reviewed studies investigating the association between personality traits and childhood obesity. Our results suggested that Conscientiousness tended to be negatively associated with the risk of childhood obesity, while consistent patterns were not shown in other personality traits of Extraversion, Agreeableness, Neuroticism, and Openness.

### Interpretation of study findings

We further explored the underlying mechanisms of the associations of personality traits with childhood obesity. We hypothesized that variables may moderate, mediate, or confound the associations (**Figure 13**). By definition, moderation refers to that the size of association differs across different levels of the moderators (i.e., age, gender); mediation refers to that personality traits indirectly influence the risk of childhood obesity via the direct influence on the mediators (i.e., eating and physical behavior, mental illness); confounding refers to a common cause of the exposure (i.e., personality traits) and the outcome (i.e., childhood obesity).

**Figure 13:**
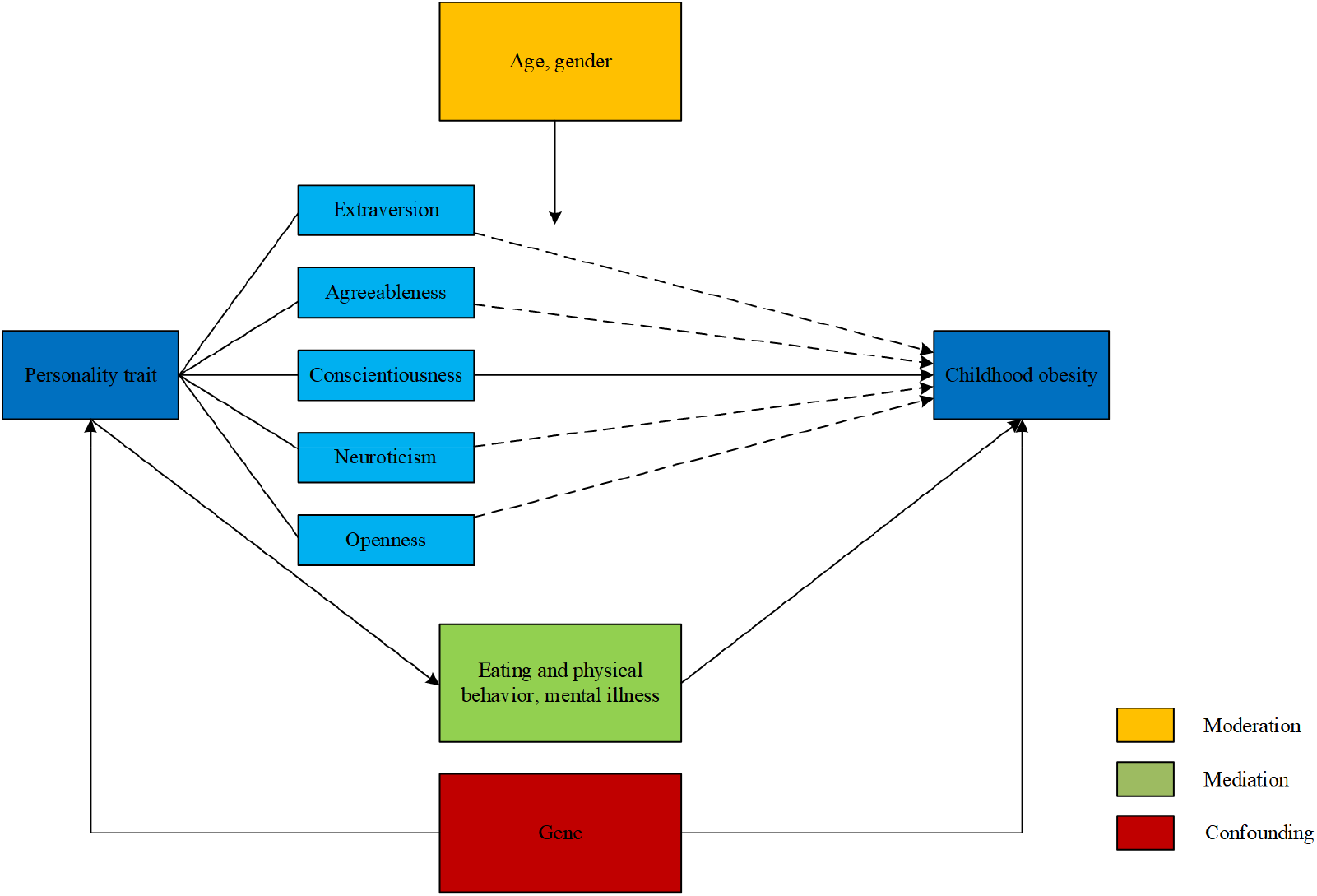
The underlying mechanisms of the association between personality traits and childhood obesity

#### Moderation

Several studies found that gender and age may moderate the associations of personality traits with childhood obesity. Vollrath et al. found girls who were less agreeable, less conscientious, and more neurotic were more likely to be overweight or obese, while similar associations were not observed in boys.^36^ Rosa et al found Agreeableness and Extraversion played a significant role in predicting BMI during the preadolescence, while other studies showed the associations of Conscientiousness with BMI in both older or younger age groups.^9,14,34^ However, most studies didn’t focus on the moderation of gender and age on the associations of personality traits with childhood obesity. Future studies should take moderation effect into consideration if the sample size permits.

#### Mediation

Diet and physical activity behaviors might mediate the associations between personality traits and childhood obesity. Vollrath et al. found that girls with lower Conscientiousness tend to consume more sugar-sweetened drinks; by contrast, more Conscientious children, particularly boys, consumed more fruits and vegetables.^36^ Another study found higher Conscientiousness was associated with lower satiety responsiveness (responding to inner cues of satiety) and food responsiveness (attracted to food cues). A meta-analytic review showed that Conscientiousness was related to eating disorders in adolescents.^37^ Several facets of Conscientiousness were also closely related to diet and physical activity behaviors. For example, concentration and perseverance correlated with lower food responsiveness and emotional overeating (eating more when feeling upset), while Impulsiveness was associated with overeating.^18,38^ Self-control was also shown to be related to reduced consumption of fast food, sugar-sweetened beverages, less sedentary behavior, and more physical activity.^26,37,39^

Mental health may also mediate the association of personality traits with childhood obesity. A meta-analysis showed high neuroticism, low conscientiousness, and low Extraversion were related to mental illnesses such as depression and anxiety and a prospective study found depression was positively associated with obesity.^40,41^

#### Confounding

It is also possible that the associations between personality traits and childhood obesity were confounded by other variables such as genes. A functional polymorphism in the promoter region of the human serotonin transporter gene (SLC6A4) is associated with several dimensions of Neuroticism.^42^ Manco et al. found some evidence for the role of SLC6A4 gene in measures of childhood obesity and Sookoian et al. found the S allele of the SLC6A4 promoter variant is associated with overweight, being an independent genetic risk factor for obesity in adolescents.^43,44^ Besides, variation in a dopaminergic gene (DRD2) is confirmed to predict Extraversion while a study by Comings et al. found 4 haplotype was in linkage disequilibrium with allelic variants of the DRD2 gene that play a major role in risk of obesity.^45,46^ A retrospective cohort study in Chinese children and adolescents with obesity also found that DRD2 rs2075654 is related to long-term remission of obesity.^47^ Similarly, the polymorphism of the brain-derived neurotrophic factor (BDNF) Val66Met is associated with Conscientiousness and obesity in adolescents.^48,49^

### Limitations and strengths

Our study is the first to comprehensively summarize the whole body of evidence about the association of personality traits with obesity in the children population. Furthermore, this review explores three possible ways (moderation, mediation, confounding) underlying the link of personality traits with the risk of childhood obesity. However, there are some limitations. We classified self-control, conformity, and persistence into other personality traits as the study authors have not clearly elaborated it under the FFM framework. It is worth noting that some previous studies considered these as facets of Conscientiousness.^22,50^ As studies we included were all written in English or Chinese, some studies written in other languages were excluded from this review. Additionally, studies that have been published tend to report significant associations between personality traits and childhood obesity, which may make our review at risk of publication bias.

## Conclusion

To conclude, studies about the associations of personality traits with childhood obesity are still in its scarcity. Although Conscientiousness has been shown a consistent association with a reduced risk of childhood obesity, more research is needed to elucidate whether the other four dimensions of personality traits also play a role in childhood obesity. Our review called for more rigorous studies in this field to uncover the causal associations of personality with obesity as well as the underlying mechanisms in the children population. The findings will be illuminating in guiding future interventions to design personality-related measures to achieve the aim of early prevention of obesity.

## Data Availability

All data produced in the present study are available upon reasonable request to the authors.

## Acknowledgements

The study was supported by National Natural Science Foundation of China (81903433). We would like to thank National Natural Science Foundation of China for its financial support.

## Abbreviations

BMI: body mass index
SMD: standardized mean difference
FFM: five-factor model
W: waist circumference
OR: odds ratio
NOS: Newcastle-Ottawa Scale

