## Supporting Information for "The association of personality traits with childhood obesity: a systematic review and meta-analysis"

**Search strategy**

PubMed: ((((‘obesity’[MeSH Terms] OR ‘overweight’[MeSH Terms]) OR ‘obese’[Title/Abstract]) OR ‘bariatric’[Title/Abstract]) OR ‘weight loss’[Title/Abstract]) OR ‘binge eating’[Title/Abstract] AND ‘personality’[Title/Abstract] AND child*[Title/Abstract]

Embase: ((((‘obesity’/exp OR ‘overweight’/exp) OR obese:ti,ab,kw) OR bariatric:ti,ab,kw) OR ‘weight loss’:ti,ab,kw) OR ‘binge eating’:ti,ab,kw AND personality:ti,ab,kw AND child*:ti,ab,kw

PsycINFO: (MA (obesity or overweight) OR TI (obese or bariatric or weight loss or binge eating) OR AB (obese or bariatric or weight loss or binge eating) OR KW (obese or bariatric or weight loss or binge eating)) AND (TI personality OR AB personality OR KW personality) AND (TI child* OR AB child* OR KW child*)

Cochrane Central Register of Controlled Trials: (((([mh“obesity”] OR [mh“overweight”]) OR obese:ti,ab,kw) OR bariatric:ti,ab,kw) OR “weight loss”:ti,ab,kw) OR “binge eating”:ti,ab,kw AND personality:ti,ab,kw AND children:ti,ab,kw
